# A Combined Predictive and Causal Approach for Neighborhood-Level Diabetes Detection

**DOI:** 10.1101/2025.02.28.25323125

**Authors:** Mohammad Noaeen, Amirhosein Rostami, Ibrahim Ghanem, Olli Saarela, Karim Keshavjee, Jeffrey R. Brook, Zahra Shakeri

**Affiliations:** Dalla Lana School of Public Health, University of Toronto, Toronto, Ontario, Canada; Institute of Health Policy, Management and Evaluation, University of Toronto, Toronto, Ontario, Canada; Department of Geography, Geomatics and Environment, University of Toronto Mississauga, Ontario, Canada; Department of Mineral and Civil Engineering, University of Toronto, Toronto, Ontario, Canada; Research Institute, The Hospital for Sick Children, Toronto, Ontario, Canada; Faculty of Information, University of Toronto, Toronto, Ontario, Canada; Schwartz Reisman Institute, University of Toronto, Toronto, Ontario, Canada

**Keywords:** machine learning, causal inference, diabetes, neighborhood-level risk, public health

## Abstract

**Objective:** Develop a neighborhood-level framework using machine learning and causal inference to identify socioeconomic and behavioral drivers of Type 2 diabetes for targeted public health interventions.

**Materials and Methods:** Data from 1,149 Census Tracts in Toronto were integrated, linking demographic, health, and marginalization indices. Seven machine learning models classified neighborhoods with high diabetes prevalence. Feature engineering mitigated skewness and correlation, while Causal Forests estimated the Conditional Average Treatment Effect (CATE, *τ*) for predictors such as work stress, smoking, and mental health.

**Results:** Predictive models achieved over 90% recall and high AUC metrics on both test and external validation datasets. Key predictors included obesity, overweight status, physical activity, and log-transformed median age. Causal analysis further indicated that elevated work stress (*τ* = 0.312) and daily smoking (*τ* = 0.155) increased diabetes risk, while stronger mental health (*τ ≈ −*1.1) was protective.

**Discussion:** While genetic and clinical factors often dominate the conversation on diabetes, data is often restricted to confirmed diagnoses or not readily available for prevalence analyses. Our study shows how neighborhood contexts, including walkability, stress levels, and socioeconomic differences, help drive rising disease rates. We integrated machine learning classifiers with causal inference to examine how interventions, such as active transportation and adjusted work stress, could shift diabetes risk.

**Conclusion:** This integrated method offers a blueprint for precision public health by clarifying how modifiable neighborhood factors affect diabetes risk. It can help tailor interventions to community needs and is applicable to other areas facing similar chronic disease challenges.

## INTRODUCTION

Type 2 diabetes affects about 422 million people world-wide (5.2% of the global population), according to the World Health Organization ^1^. In Canada, over three million individuals (8.9% of the population) have been diagnosed with diabetes, and 6.1% of adults aged 20 to 79 are prediabetic ^2^. Prevalence is projected to rise by 3.3% each year ^2^. Although traditional perspectives emphasize biological factors, growing evidence shows that social determinants of health (SDoH) shape disease incidence and outcomes ^3–5^. SDoH cover the conditions in which people live and work, including neighborhood-level resources and stressors. Green space access, housing stability, and walkability are linked to better metabolic health ^6–9^. Socioeconomic gaps, such as differences in income, employment, and education, correlate with diabetes burden ^10,11^. Regional studies in Ontario and Finland show how these factors vary, complicating prevention efforts ^12–14^. These complexities suggest a need for methods that capture both the breadth of environmental contributors and the localized nature of risk. Neighborhood-level approaches link environmental factors to disease dynamics in ways that align with clinical priorities and social service needs ^15^. Community-level SDoH data can substitute for missing individual information and capture broader contextual influences on health behaviors, strengthening the rationale for including geographically detailed data in predictive models.

Recent machine learning (ML) advancements have improved predictive analytics and risk modeling in health research by using varied datasets to study disease occurrence and progression ^16–18^. Researchers have created ML models for multiple purposes: prognostic modeling of diabetes ^19^; predicting diabetes, prediabetes, and complications with ensemble methods ^20–22^ and deep learning ^23–26^; examining mental health comorbidities with deep neural networks ^27^ and ensemble strategies ^28^; exploring epidemiological trends using population-level modeling ^29^; and estimating time to diabetes onset with survival analysis ^30,31^. Some newer efforts have used large language models (LLMs) and generative AI for diabetes prevention and support ^32,33^, although applying them at the neighborhood level can be difficult. Traditional classification and regression algorithms often provide clearer interpretability of contextual factors, like local socioeconomic conditions, especially when data is aggregated or partial. Many diabetes-focused ML projects rely on individual-level data ^34^, but neighborhood-oriented perspectives can uncover spatial trends that standard clinical repositories may miss, particularly in urban areas with strong socioeconomic differences ^35^.

Building on neighborhood-level perspectives, conventional ML methods often detect correlations without determining whether changing a given factor would affect health outcomes ^36^. An algorithm might link green space with lower diabetes prevalence but not show if expanding green space actually reduces disease rates. Causal machine learning (Causal ML) combines statistical inference with ML approaches to address such ‘what if’ questions and provide deeper insights into possible outcomes of targeted interventions ^37–39^. This framework aids targeted public health efforts by indicating how factors like walkability or income might serve as key prevention levers. Although public health agencies track patient-level diabetes data, they typically focus on confirmed cases, overlooking psychosocial or environmental factors at finer scales. ML-based methods integrate these variables to identify high-risk neighborhoods and guide localized interventions that account for underlying social and environmental conditions.

Our study applies both standard ML and Causal ML methods to forecast neighborhood-level diabetes prevalence in the Toronto Census Metropolitan Area (Toronto CMA, population 6.2 million) and the City of Toronto ^40^. The city’s cultural and ethnic variety broadens the reach of these insights. This combined approach supports precision public health by finding hotspots for intervention and adapting risk reduction strategies to local needs. Clinicians, policymakers, and urban planners can apply these findings to shape resource allocation plans that consider socioeconomic and environmental conditions. The proposed framework links predictive results to real-world impact, illustrating how SDoH vary among communities and guiding targeted prevention and improved care.

## MATERIALS AND METHODS

Figure 1 outlines the pipeline for identifying neighborhoods with high type 2 diabetes prevalence in the Toronto CMA. The process encompasses data collection, feature engineering, predictive modeling with seven ML algorithms, and interpretative analyses supported by sensitivity checks, SHapley Additive exPlanations (SHAP) analysis, and causal ML to explore potential causal pathways. We define our dataset as 𝒟 = {(*x*_1_, *y*_1_), (*x*_2_, *y*_2_), …, (*x*_*n*_, *y*_*n*_)}, where each neighborhood *i* has a feature vector *x*_*i*_ ∈ ℝ^*m*^ (with *m* total features) and a binary label *y*_*i*_ indicating whether the neighborhood lies in the top quintile of diabetes prevalence (10.5% to 23.8%). A model *M* is trained to predict high-prevalence status, so 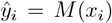 accurately flags neighborhoods at greater risk.

**Figure 1.**
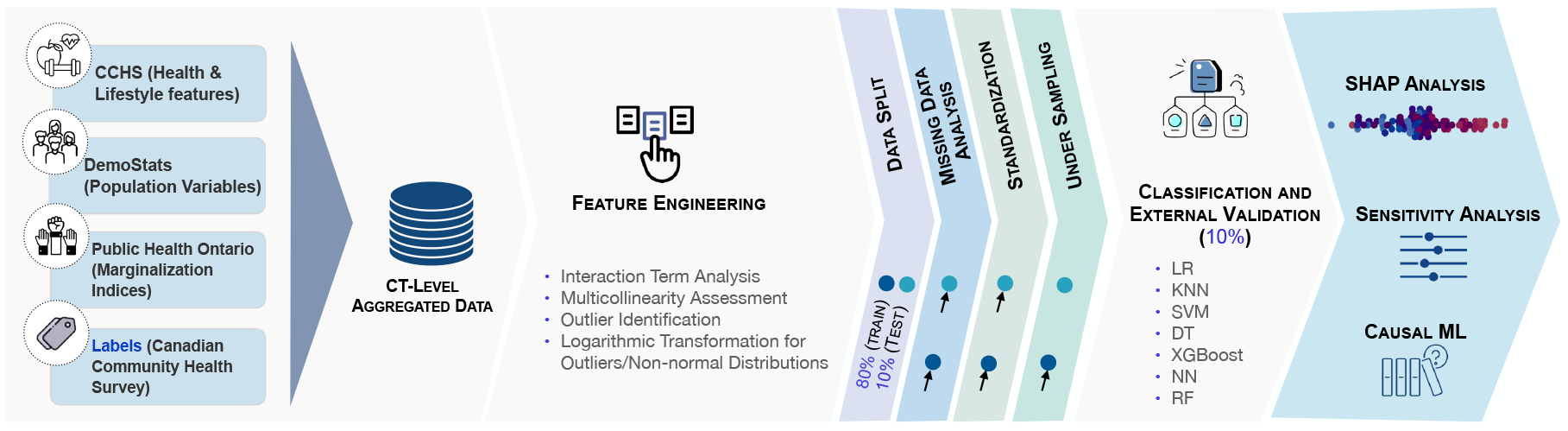
Overview of the employed ML and Causal ML pipeline. The workflow combines data from multiple sources, addresses interactions and class imbalance, then trains seven ML algorithms (LR, KNN, SVM, DT, XGBoost, NN, RF). SHAP, sensitivity analysis, and Causal ML clarify model outputs and suggest potential neighborhood interventions.

### Dataset Overview and Feature Engineering

#### Dataset Overview

Census tracts (CTs) defined by Statistics Canada outline neighborhoods with comparable socioeconomic and living conditions ^41^. We chose predictors based on prior studies linking health, socioeconomic, lifestyle, and marginalization indicators to diabetes prevalence ^4,6,8,9,11^. We obtained type 2 diabetes incidence from the 2022 Canadian Community Health Survey (CCHS) ^42^, which collects data on Canadians aged 12 and older in 121 health regions, covering 98% of the population. The remaining 2% live on Indian Reserves, Crown Lands, or remote areas not covered due to limited infrastructure and practical barriers. Figure 2a displays 1,149 neighborhoods in the Toronto CMA, of which 230 (20.0%) are high-prevalence. A neighborhood was labeled high-prevalence if its diabetes rate exceeded the 80th percentile (10.5%). Figure 2b shows values up to 23.8%, so 10.5% to 23.8% falls into this high-prevalence band. This binary categorization highlights areas with elevated risk.

**Figure 2.**
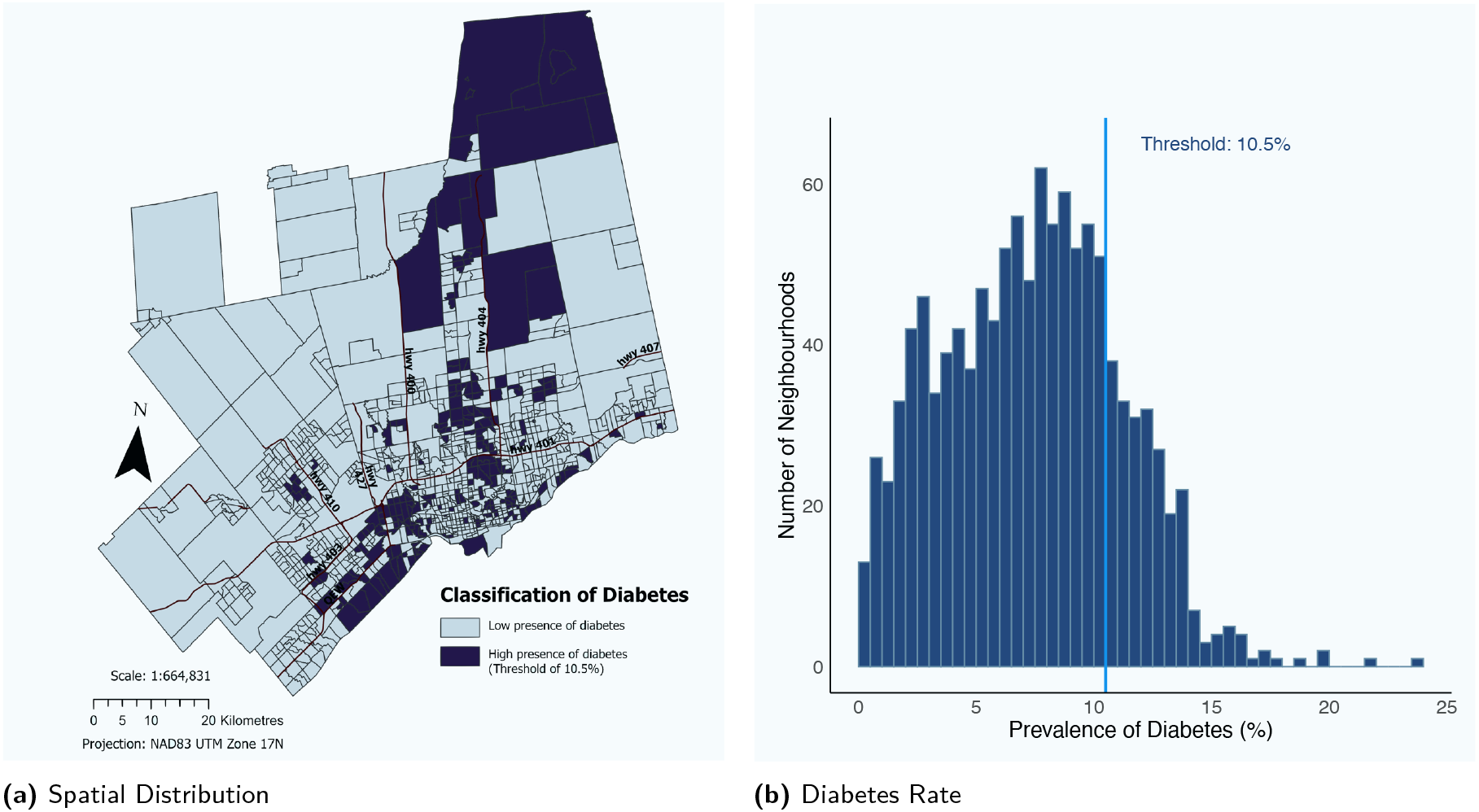
Geographic Distribution and Prevalence Rates of Type 2 Diabetes in the Toronto CMA. (a) Map of neighborhoods classified as high-prevalence (10.5%–23.8%) or low-prevalence. (b) Histogram showing the distribution of Type 2 diabetes prevalence rates across 1,149 neighborhoods in the region.

#### Data Exploration and Feature Engineering

The dataset assigned a unique identifier to each of the 1,149 CTs in the Toronto CMA. Three main sources provided neighborhood variables: health and lifestyle measures from the 2022 CCHS, demographic and socioeconomic data from Environics Analytics’ DemoStats ^43^, and 2021 Marginalization Indices from the MAP Centre for Urban Health Solutions ^44^. DemoStats offers more than 750 demographic indicators derived from census data, economic data, and immigration information, accessible across various administrative divisions ^45^. The Marginalization Indices measure neighborhood-level factors, such as income or housing inequalities, using factor scores and quintile ranges ^46^. Data came from SimplyAnalytics ^47^ and Public Health Ontario ^48^, leading to 26 predictive features summarized in Table 1.

**Table 1.** Features were sourced from the Canadian Community Health Survey (CCHS), DemoStats, and Public Health Ontario, representing population health, demographics, and socio-economic status.

| Feature | Description |
| --- | --- |
| **Canadian Community Health Survey** |  |
| Mental health score | Perceived mental health: Higher = better |
| Mental health binary * | 0 = poor, 1 = good (avg score cutoff) |
| Regular alcohol drinkers rate | % regular alcohol drinkers |
| Daily smokers rate | % daily smokers |
| Food insecurity score | Food insecurity: Higher = more |
| Food insecurity binary* | 0 = low, 1 = high (avg score cutoff) |
| Work stress score | Perceived work stress: Higher = more |
| Work stress binary* | 0 = low, 1 = high (avg score cutoff) |
| Rate of active population | % physically active (18+) |
| Active transportation rate | % using active transport (18+) |
| Obese rate | % obese population |
| Overweight rate | % overweight population |
| **DemoStats** |  |
| High education rate | % with diploma/degree (25-64) |
| Average income | Avg household income |
| Unemployment rate | % unemployed (15+) |
| Median age | Median population age |
| Recent immigrant rate | % immigrants (2017-2022) |
| Visible minority rate | % visible minorities |
| Visible minority binary* | 0 = <25%, 1 = >25% |
| Rented rate | % rented dwellings |
| **Public Health Ontario** |  |
| Ethnic concentration index | % recent immigrants & visible minorities |
| Ethnic concentration quintiles | Ethnic concentration quintiles |
| Residential instability index | Residential density & family structure |
| Residential instability quintiles | Residential instability quintiles |
| Material deprivation index | Access to basic needs |
| Material deprivation quintiles | Material deprivation quintiles |

\* Feature calculated by the authors using the raw feature

We applied standard procedures like one-hot encoding for categorical variables and created interaction terms to capture more complex relationships. For example, an ‘instability deprivation’ variable combined material deprivation and residential instability quintiles ^44^, reflecting neighborhoods facing both social and economic difficulties. Log transformations addressed skewness in factors such as median age, and we reviewed thresholds to exclude collinear attributes exceeding |*r*| *≥* 0.7.

The dataset was split into training (80%), test (10%), and external validation (10%) sets, with feature scaling confined to the training subset before model fitting. Given the 20% positive-class rate, we applied Random Under-Sampling (RUS) solely to the training data, reducing it from 919 to 340, while keeping the test and validation sets unchanged. We designated neighborhoods in Old Toronto (along Lake Ontario) and Brampton for external validation in the Toronto CMA and Toronto, respectively. Prevalence of type 2 diabetes in Peel Region, which includes Brampton, is notably higher than the Ontario average ^49^. Both validation samples contained a mix of high and low prevalence neighborhoods, ensuring a diverse basis for model evaluation. Five-fold crossvalidation helped refine hyperparameter tuning and feature selection, improving model consistency.

### Predictive Modeling

A hypothesis class ℋ with seven ML architectures was chosen to reflect varied learning capacities and interpretability: Logistic Regression (LR), K-Nearest Neighbors (KNN), Neural Networks (NN), Support Vector Machines (SVM), Decision Trees (DT), Random Forests (RF), and Extreme Gradient Boosting (XG-Boost). These algorithms differ in complexity, feature handling, and parameterization, allowing an in-depth examination of neighborhood-level heterogeneity. Each model learns a decision rule *h ∈*ℋ to minimize a loss function on the training set, with hyperparameters tuned by five-fold cross-validation.

We evaluated each model’s performance by accuracy, precision, recall, F1, and area under the curve (AUC) to provide a thorough assessment of predictive ability. Recall was prioritized to ensure effective identification of high-prevalence neighborhoods. We selected optimal hyperparameters via grid search with five-fold crossvalidation, and used Recursive Feature Elimination with Cross-Validation (RFECV) for LR, NN, DT, RF, and XGBoost to remove redundant predictors. We summarized results with confusion matrices and ROC curves for training and testing. The best models were tested on geographically distinct external subsets, and SHAP provided both localized and global interpretations of model outcomes while measuring each feature’s contribution ^50^. Table 2 lists the final feature sets and hyperparameters identified through grid search. These features, obesity and overweight rates, active transportation and population activity rates, daily smoking rates, visible minority status, and recent immigration patterns, show strong links to diabetes prevalence. Their repeated appearance across models suggests their importance for populationlevel interventions.

**Table 2.** Summary of final feature sets (determined by recursive feature elimination with cross-validation) and model parameters (determined by grid search with 5-fold cross-validation) across the seven machine learning models.

| Model | # of features | Optimal set of features | Model parameters |
| --- | --- | --- | --- |
| LR | 5 | Obese rate, Overweight rate, Active transportation rate, Rate of active population, Log median age | $C^{[1]}$: 0.3, *penalty*: l1, *solver*: liblinear |
| KNN | 5<sup>[2]</sup> | Obese rate, Overweight rate, Active transportation rate, Rate of active population, Log median age | Optimal $k$ (using Elbow method): 6, *algorithm*: auto, *metric*: cosine and *weights*: uniform |
| NN<sup>[3]</sup> | 7 | Obese rate, Overweight rate, Active transportation rate, Rate of active population, Log median age, Visible minority rate, Daily smokers rate | *activation*: tanh, *hidden_layer_sizes*: (128, 64), *solver*: lbfgs |
| SVM | 5<sup>[2]</sup> | Obese rate, Overweight rate, Active transportation rate, Rate of active population, Log median age | $C^{[1]}$: 10, *gamma*: 0.1, and *kernel*: rbf<sup>[4]</sup> |
| DT | 4 | Obese rate, Overweight rate, and Log median age, Visible minority rate | *max_depth*: 20, *min_samples_leaf*: 5, *max_features*: square root, and *splitter*: best |
| RF | 7 | Obese rate, Overweight rate, Active transportation rate, Rate of active population, Log median age, Visible minority rate, Daily smokers rate | *max_depth*: 10, *min_samples_leaf*: 3, *max_features*: log2, and *n_estimators*: 150 |
| XGBoost | 6 | Obese rate, Overweight rate, Rate of active population, Visible minority rate, Daily smokers rate, Recent immigrant rate | *max_depth*: 5, *min_samples_leaf*: 10, *n_estimators*: 150, and *learning_rate*: 0.1 |

<sup>[1]</sup> Regularization Parameter, <sup>[2]</sup> Same optimal feature set as LR model, <sup>[3]</sup> Multi-Layer Perceptron, <sup>[4]</sup> Radial Basis Function

All machine learning models in this project were built with `Python` 3.11 and `Scikit-learn` 1.6.1, and are available on GitHub^1^, and an interactive version is hosted via the project dashboard^2^. We exported them with `PyMilo`^3^, which saves each model’s settings in a secure, non-executable JSON file to enable cross-platform compatibility and address safety concerns associated with binary formats, an important consideration in healthcare.

### Causal Inference through Causal ML: Causal Forests

Causal ML goes beyond correlation-based predictions by examining how shifting a specific factor might change an outcome ^51,52^. Although randomized controlled trials are the gold standard for causal inference, they can be impractical at the neighborhood scale, where randomizing entire communities presents logistical and ethical concerns. Causal methods use observational data to approximate such scenarios by adjusting for confounding and comparing interventions across similar areas, supporting policy decisions on how altering a feature could influence disease prevalence.

In this study, we use the same dataset 𝒟 = {(*x*_*i*_, *y*_*i*_)} of neighborhoods, but introduce a treatment variable *t*_*i*_ ∈ {0, 1} that indicates whether a specific intervention or environmental factor is present. Potential outcomes *Y* (0) and *Y* (1) are the diabetes prevalence under control and treatment conditions, respectively. We define the Conditional Average Treatment Effect (CATE) as *τ* (*x*) = 𝔼 [*Y* (1) *− Y* (0) | *X* = *x*], where 𝔼 is the expectation, representing the average difference in outcomes between treatment and control for neighborhoods sharing covariates *x*.

We used Microsoft Research’s `EconML` library to implement Causal Forests, an adaptation of Generalized Random Forests that captures variation in treatment effects ^53^. These forests split the feature space so that neighborhoods with distinct treatment effects fall into different leaves, potentially identifying populations that would benefit more from interventions like healthier food access or green space. Estimating 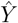 (1) and 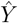 (0) clarifies how interventions might shift local diabetes rates, showing whether some neighborhoods respond more strongly to changes in walkability, income assistance, or other features. This approach supports more precise targeting of public health measures through the simulation of counterfactual scenarios.

A total of 10,000 trees were used to improve estimate stability, and parameters were tuned with 10-fold crossvalidation. We used LassoCV as the base estimator for both outcome and treatment models, imposing a regularization penalty that helps avoid overfitting by promoting sparse solutions.

## RESULTS

Table 3 compares seven ML algorithms for detecting neighborhood-level diabetes prevalence in the Toronto CMA and the City of Toronto, focusing on accuracy, precision, recall, and F1. SVM and NN stood out as leading methods; SVM correctly identified all high-prevalence neighborhoods (*y*_*i*_ = 1) and achieved 100% recall. The NN (multi-layer perceptron) followed, recording 95% recall on the test set and 78% on the external validation set. Figure 3 shows that SVM reached an AUC of 0.96 on the test dataset, the top result among the models, while NN reached an AUC of 0.94 on the test set and 0.86 in external validation.

**Table 3.** Summary of performance metrics for each model, including accuracy, precision, recall, and F1 score, on test (T) and external validation (E) datasets for the Toronto CMA and the City of Toronto.

| Model | Toronto CMA |  |  |  |  |  |  |  | City of Toronto |  |  |  |  |  |  |  |
| --- | --- | --- | --- | --- | --- | --- | --- | --- | --- | --- | --- | --- | --- | --- | --- | --- |
|  | Acc |  | Prec |  | Recall |  | F1 |  | Acc |  | Prec |  | Recall |  | F1 |  |
|  | T | E | T | E | T | E | T | E | T | E | T | E | T | E | T | E |
| LR | 0.82 | 0.82 | 0.51 | 0.44 | 0.90 | 0.67 | 0.66 | 0.53 | 0.81 | 0.80 | 0.40 | 0.40 | 1.00 | 0.73 | 0.54 | 0.51 |
| KNN | 0.86 | 0.87 | 0.58 | 0.55 | 0.90 | 0.89 | 0.70 | 0.67 | 0.83 | 0.84 | 0.41 | 0.43 | 0.93 | 0.91 | 0.56 | 0.59 |
| NN | 0.87 | 0.89 | 0.59 | 0.61 | 0.95 | 0.78 | 0.73 | 0.68 | 0.82 | 0.84 | 0.45 | 0.45 | 1.00 | 0.90 | 0.62 | 0.61 |
| SVM | 0.86 | 0.85 | 0.57 | 0.52 | 1.00 | 0.94 | 0.72 | 0.67 | 0.83 | 0.77 | 0.45 | 0.41 | 1.00 | 0.91 | 0.62 | 0.62 |
| DT | 0.79 | 0.83 | 0.45 | 0.48 | 0.62 | 0.83 | 0.52 | 0.61 | 0.84 | 0.84 | 0.38 | 0.45 | 0.74 | 0.85 | 0.50 | 0.56 |
| RF | 0.86 | 0.82 | 0.58 | 0.47 | 0.86 | 0.89 | 0.69 | 0.62 | 0.79 | 0.74 | 0.37 | 0.37 | 0.85 | 0.86 | 0.51 | 0.51 |
| XGBoost | 0.75 | 0.75 | 0.41 | 0.38 | 0.71 | 0.83 | 0.52 | 0.54 | 0.74 | 0.78 | 0.39 | 0.39 | 0.63 | 0.78 | 0.51 | 0.51 |

**Figure 3.**
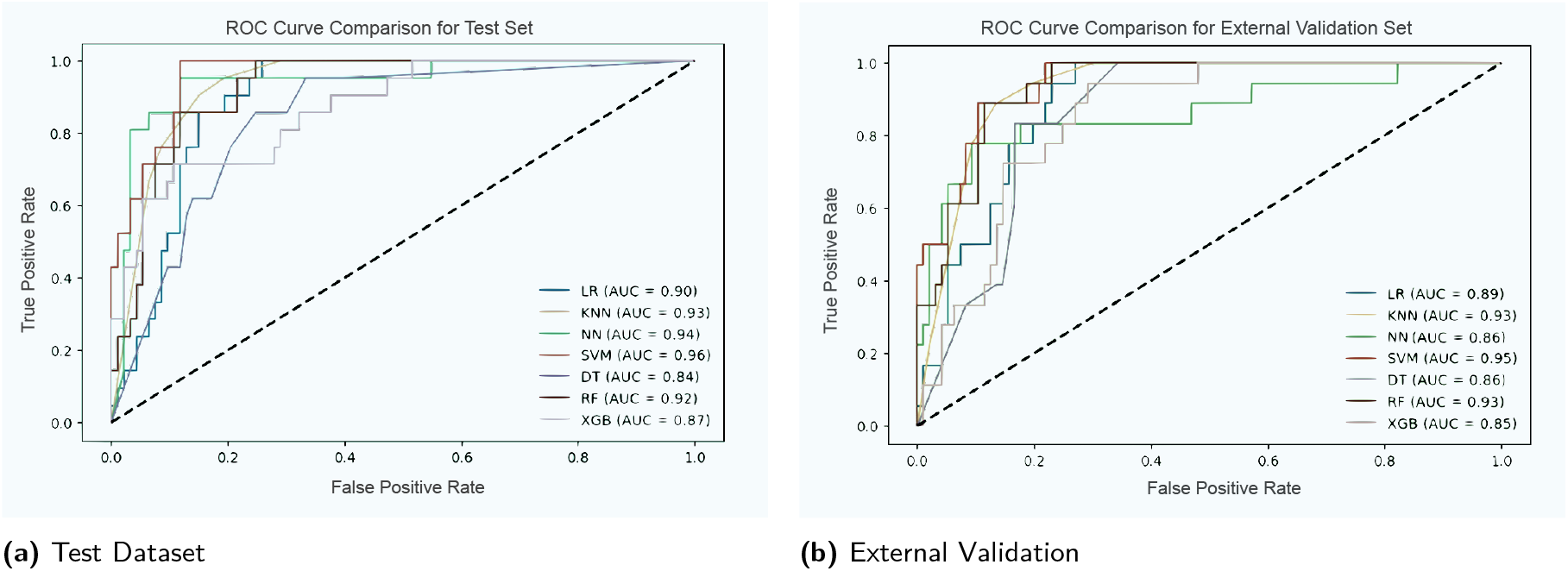
ROC Curve Comparison for Diabetes Prevalence Prediction. Each curve shows the true positive rate (TPR) and false positive rate (FPRs) across different thresholds. Higher TPRs and lower FPRs shift the curve further away from the diagonal line, which represents random guessing (TPR = FPR). The SVM model outperforms other methods, achieving the highest AUC of 0.96 on the test set and 0.95 on the external validation set.

Obesity, overweight status, and active transportation were especially influential in distinguishing neighborhoods with higher diabetes prevalence, aligning with evidence that weight gain and physical inactivity elevate insulin resistance ^4,6^. SVM and NN captured non-linear interactions in these inputs by encoding them in richer feature spaces: the SVM applied a Radial Basis Function kernel to isolate boundaries influenced by socioeconomic and environmental factors, and the NN used multiple hidden layers to interpret interactions among variables such as education, walkability, and obesity. Other algorithms in ℋ (e.g., DT, RF, and XGBoost) also model non-linearities, but the SVM and NN demonstrated superior accuracy and recall on this dataset.

During data preparation, an initial SVM setup achieved 88% recall. After applying logarithmic scaling and removing highly correlated attributes, recall rose to 100%. This change suggests that correcting skewed distributions and pruning redundant data helped clarify the classifier’s decisions, making it more capable of identifying high-prevalence neighborhoods. Although a 100% recall may raise questions of overprediction, the SVM retained balanced precision and F1 scores, indicating it did not classify all neighborhoods as 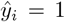. These results suggest the model accurately captured truly high-prevalence neighborhoods without markedly boosting the false-positive rate.

Using the RFECV procedure, five leading predictors emerged for the SVM and LR models: obesity, overweight, active transportation, rate of active population, and log-transformed median age (Table 2). These factors align closely with local living conditions and health behaviors, emphasizing the usefulness of blending individual lifestyle indicators with environmental cues when modeling diabetes risk.

### Geographical Applicability and Model Scalability

The Toronto CMA covers a larger and more diverse region, while the City of Toronto forms its urban core with distinct demographics and fewer wide-ranging socioeconomic contrasts. Applying the trained SVM model to the City of Toronto lowered precision (from 57% to 45% in the test dataset and from 52% to 41% in the external validation dataset), while recall stayed at 100% in the test dataset but dipped from 94% to 91% in external validation (Table 3). One explanation is that the City of Toronto has a narrower range of certain predictors, leading the classifier to label more neighborhoods as high-prevalence. In that subset, 20% of neighborhoods are positive, so a 45% precision still doubles the base rate. From a public health view, consistently identifying areas of greater prevalence may be preferable, even if it requires extra follow-up to rule out false positives.

### Causal Analysis of Diabetes Risk Factors

We compared three sets of predictors to capture various neighborhood dynamics: (1) the base SVM set (log-transformed median age, obesity, overweight, active transportation, and active population rate), (2) an extended set that added mental health score, visible minority rate, work stress, and daily smokers rate, and (3) a broader range of attributes (Table 4). The extended set included psychosocial factors such as stress and daily smoking, while managing model complexity.

**Table 4.**
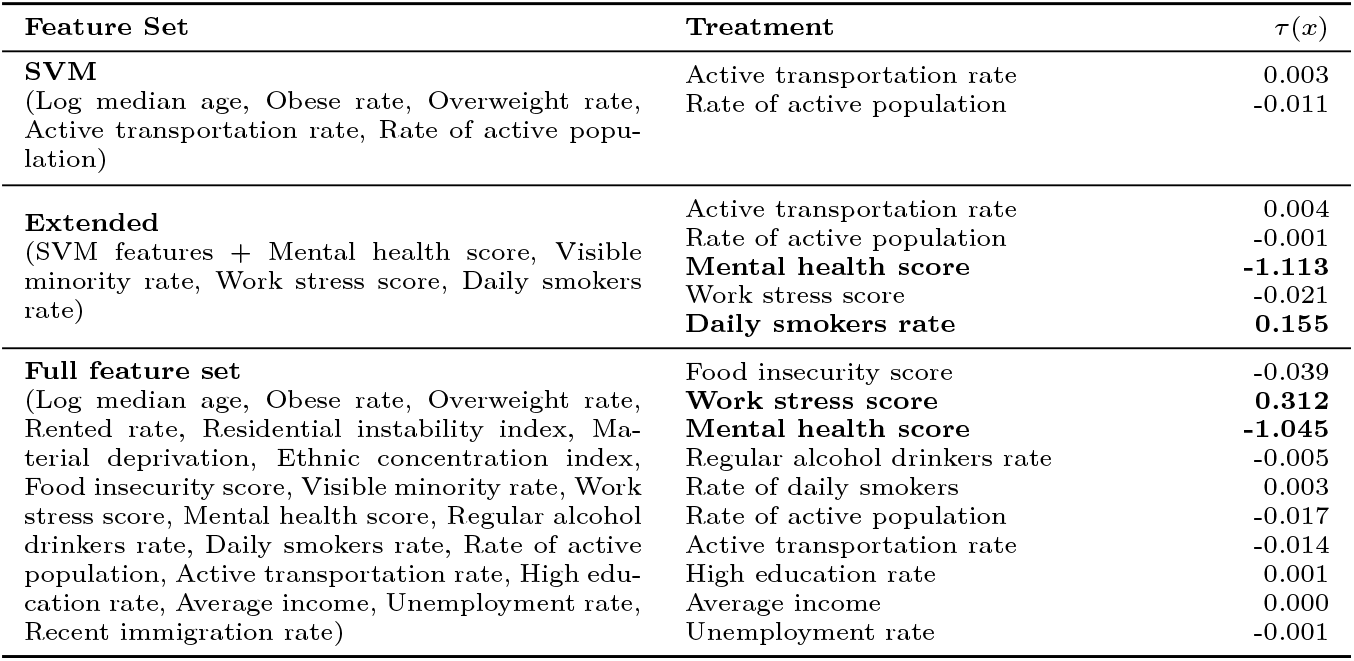
Summary of Causal Forest results for each feature set, reporting the estimated *τ* (*x*) for selected treatments in neighborhood-level diabetes prevalence.

We used Causal Forests to estimate *τ* (*x*) (i.e., the CATE) and explore how socioeconomic, environmental, and behavioral variables relate to neighborhood-level diabetes prevalence. Some attributes (log-transformed median age, obesity, overweight, visible minority rate, and recent immigrant rate) were excluded as treatment variables because adjusting them in real life typically involves overlapping efforts. For example, reducing obesity at scale requires integrated policies, nutrition guidance, and community resources, and recent immigration status may act as a confounder linked to multiple factors like income and education.

With the SVM’s primary predictive features (logtransformed median age, obesity, overweight, active transportation, and active population rate), the Causal Forest estimates showed minimal direct effects for active transportation *τ* (*x*) = 0.003 and active population rate *τ* (*x*) = *−* 0.011. This outcome does not imply that active transportation lacks value, only that within our dataset and approach, it did not exhibit a strong causal influence. When we extended the SVM feature set (adding mental health score, visible minority rate, work stress, and daily smokers rate), mental health produced a meaningful protective effect *τ* (*x*) = *−* 1.113, and daily smoking aligned with known metabolic risks *τ* (*x*) = 0.155.

In the broader set, work stress carried a modest positive coefficient *τ* (*x*) = 0.312, while mental health remained strongly negative *τ* (*x*) = *−* 1.045. Food insecurity showed a slight negative coefficient *τ* (*x*) = *−* 0.039, possibly reflecting a complex mix of household factors. Alcohol use, unemployment, and recent immigration each had negligible effects, implying that other psychosocial and environmental features play a larger role in neighborhood-level diabetes vulnerability.

## DISCUSSION

Neighborhood-level data provided a clear edge in identifying communities with high diabetes prevalence, extending beyond insights from individual-level or heavily aggregated sources. When combined with physical activity indicators, obesity, overweight rates, and age structure emerged as important predictors, showing how built environments and lifestyle factors shape local risk ^54,55^. These findings align with prior research connecting cardiometabolic conditions to excess body weight and sedentary living, especially in urban areas ^4^. However, standardized neighborhood-level data remain limited in many health agencies, making it difficult to track socioeconomic variations in diverse populations ^56^. Systematic inclusion of local characteristics could refine resource allocation by identifying areas where interventions are most urgently needed ^57^.

In this study, we dealt with class imbalance because neighborhoods labeled *y*_*i*_ = 1 were a small portion of 𝒟. Such imbalance can bias standard ML algorithms toward majority classes ^58,59^. For *N*_maj_ representing the majority class size, *N*_min_ the minority class size, and *r* the target ratio, we picked a subset *S ⊂ N*_maj_ with |*S*| = *r* · *N*_min_. We then applied random under-sampling (RUS) to remove majority examples until reaching that ratio, improving attention to high-prevalence neighborhoods. RUS avoids generating synthetic data that could distort the complex socioeconomic patterns in our categorical and survey-based features. Methods like SMOTE ^60^ interpolate continuous features as:

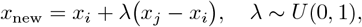

where *U* (0, 1) is the continuous uniform distribution. Although SMOTE-NC adapts this approach for mixed data, synthetic points may misrepresent certain categorical or survey-derived characteristics essential to our local context. RUS preserves authentic minority samples, aligning with our goal of maximizing recall for higher-risk neighborhoods ^61^.

We also explored probability calibration with methods such as Platt scaling or isotonic regression. Platt scaling fits a logistic function,

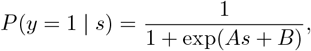

where *s* is the uncalibrated score, and (*A, B*) are parameters estimated by maximum likelihood. Isotonic regression produces a piecewise constant function that aligns scores with probabilities. These approaches can refine probability estimates but may also add complexity, especially in smaller datasets. Our primary objective was to identify neighborhoods with high prevalence rather than deliver fully calibrated probabilities, so we prioritized recall and F1 scores over precise calibration.

Thoughtful selection of neighborhood features improves model performance and guides practical public health efforts. Including factors like material deprivation and residential instability reveals both individual risk elements and broader community challenges, shaping decisions on infrastructure and resource allocation. This integrated approach links our predictive outcomes to real-world actions and can extend to other chronic diseases affected by environmental and behavioral influences, such as hypertension.

### Assessing Model Reliability and Sensitivity

We conducted a feature permutation test ^62^ to assess each predictor’s impact on a trained model *f* (x). With *M* (*f*) representing a performance metric (e.g., recall or F1), each feature *x*_*j*_ was randomly permuted to create a dataset 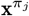. The sensitivity of *x*_*j*_ is:

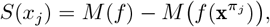

where *M*(*f* (x)) is the original score. Larger differences indicate greater feature importance ^63^. Figures 4a and 4b show that permuting `Log Median Age` decreased recall and F1 the most, suggesting its importance in detecting areas with higher diabetes prevalence. `Obesity`, `Overweight`, and `Physical Activity` followed closely, aligning with studies linking active living to lower metabolic risk in urban contexts ^64^.

**Figure 4.**
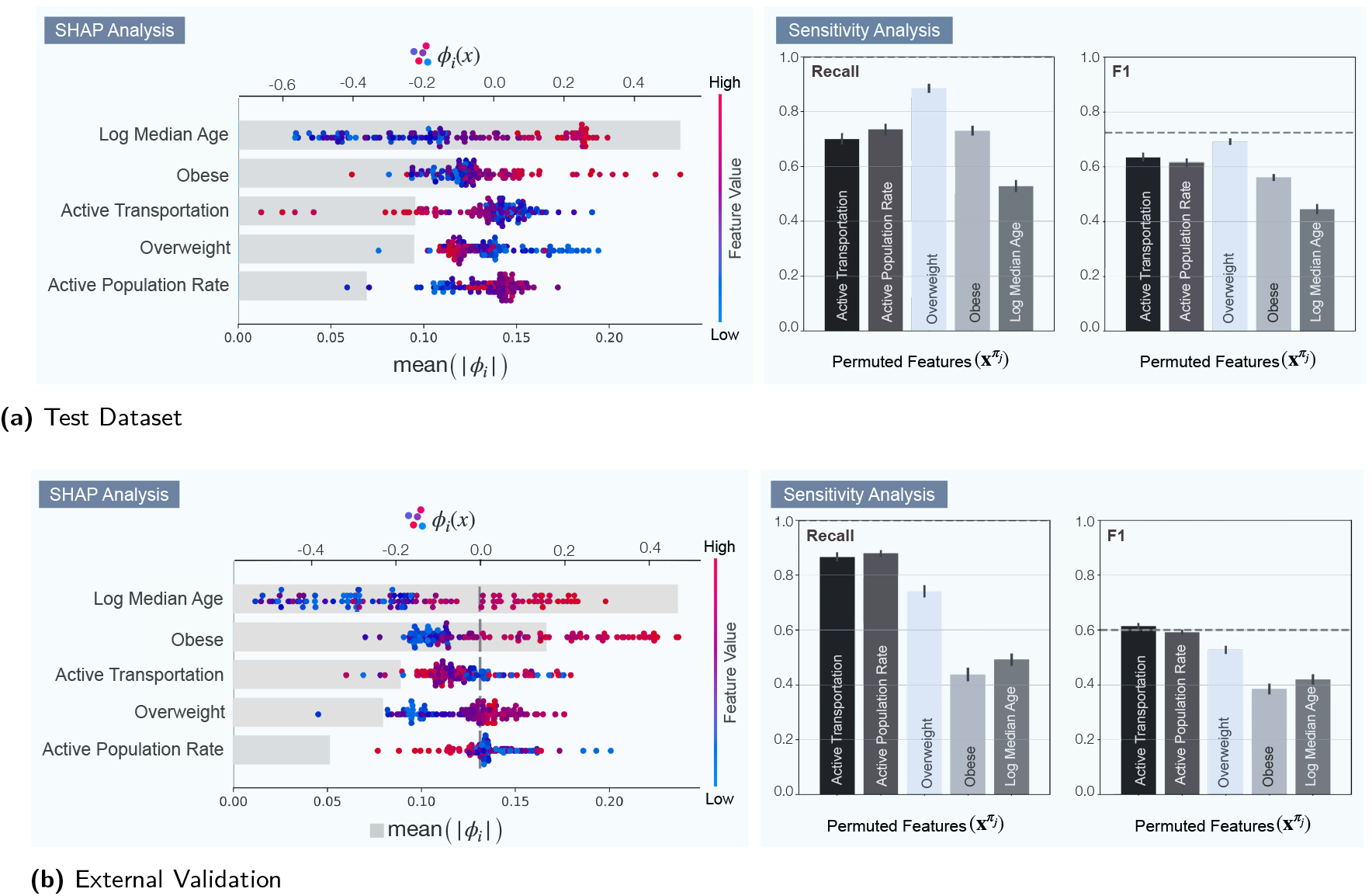
Integrated SHAP and Sensitivity Analysis Illustrating SVM Interpretability and Robustness. In each panel, a dual-axis plot displays the SHAP component: the top axis features a beeswarm plot of local Shapley values *ϕ*_*i*_(*x*) for each feature *i* in instance *x* (with a pink-to-blue gradient for low-to-high feature values; values right of the vertical line indicate positive contributions that increase the predicted outcome, and those left indicate negative contributions), while the bottom axis shows a Barnhart plot of global feature importance via mean absolute SHAP values 𝔼 [| *ϕ*_*i*_|] (with higher values signifying greater impact). Adjacent bar charts show the change in Recall and F1 metrics following feature permutation (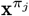, where the *j*th feature is shuffled to break its association with the target), illustrating model robustness across the Test (a) and External Validation (b) datasets.

We also performed a SHAP analysis to examine each variable’s localized contribution to the predicted outcome. SHAP values *ϕ*_*i*_(x) break down the difference between a baseline prediction and an instance’s prediction across all features ^50^:

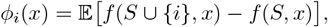

where the expectation is taken over all feature subsets *S* that exclude *i*^50^. Figures 4a and 4b provide beeswarm plots illustrating how higher values for `Log Median Age` or `Obesity` push predictions toward high diabetes prevalence (points to the right), while more active transportation drives the predicted risk downward (points to the left). The adjacent bar charts confirm that these same predictors strongly affect recall and F1 when permuted, indicating a match between the global permutation results and local SHAP attributions. This synergy suggests that focusing on these features could aid communities at higher risk ^65,66^.

#### Urban Lifestyle Interventions

Our findings show that certain aspects of urban living, especially those tied to physical activity, have a measurable link to neighborhood-level diabetes risk. Recent studies highlight the built environment’s role in shaping physical activity habits and metabolic health ^67^. Our estimated *τ* (*x*) suggests that adjustments in urban design could reduce diabetes prevalence in at-risk areas, reflecting the idea that well-planned infrastructure encourages healthier behaviors.

Evidence from major population centers suggests that streets designed for pedestrians and bicycles correlate with lower diabetes rates ^9,68^. In Toronto, debates over removing bike lanes have raised concerns about effects on daily activity and well-being ^69^. Similar outcomes in Copenhagen and Melbourne show that designs promoting active transportation align with improved metabolic indicators ^70^. Urban lifestyle interventions also offer benefits beyond diabetes reduction. Facilities supporting walking and cycling contribute to cleaner air and strengthen social connections ^71^. Policymakers may use these insights to align urban planning with public health goals, clarifying how specific design choices within built environments can reinforce healthier lifestyles.

#### Ethnocultural Influences on Diabetes

We used the same features in both the City of Toronto and Toronto CMA datasets. In the City of Toronto dataset, mental health and work stress scores were removed due to multicollinearity. An extended analysis of the Toronto CMA dataset included mental health score, work stress score, log median age, obesity, overweight, active transportation, and active population rate (Table 4). Adding these predictors led to a 2% gain in accuracy, a 6% gain in precision, and a 1% gain in F1, but a 3% drop in recall. This aligns with earlier work showing that broadening a model’s feature space boosts accuracy and precision at the cost of slight sensitivity loss ^72,73^. The trade-off arises because additional predictors introduce variability, making positive cases easier to identify, even if some borderline examples may be missed. Including mental health and work stress measures enhances the model’s ability to capture local sociocultural factors in diverse areas like the Toronto CMA, where these variables affect diabetes risk.

In our analysis, visible minority rates were a key predictor of neighborhood-level diabetes prevalence. To explore this relationship, we used a binned average apneighborhoods in bin and denotes diabetes status proach: 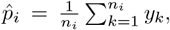,, where *n*_*i*_ is the number of for neighborhood *k*. The fitted line in Figure 5a shows the mean prevalence per bin, with a 95% confidence interval given by ^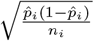^. We observed an inverse relationship between visible minority rates and diabetes prevalence, indicating that higher proportions of visible minorities tended to coincide with lower average diabetes prevalence. This pattern supports earlier findings on the links between neighborhood racial composition, residential segregation, and diabetes disparities ^74,75^. It may also suggest that neighborhoods with more visible minorities sustain traditional dietary habits and other culturally protective practices ^76^, and that the ‘healthy immigrant effect’ adds further protection since many newcomers arrive in better health due to selective immigration policies ^77^. In Toronto, where more than 50% of residents are immigrants ^78^, these observations show how neighborhood demographics can influence diabetes burden.

**Figure 5.**
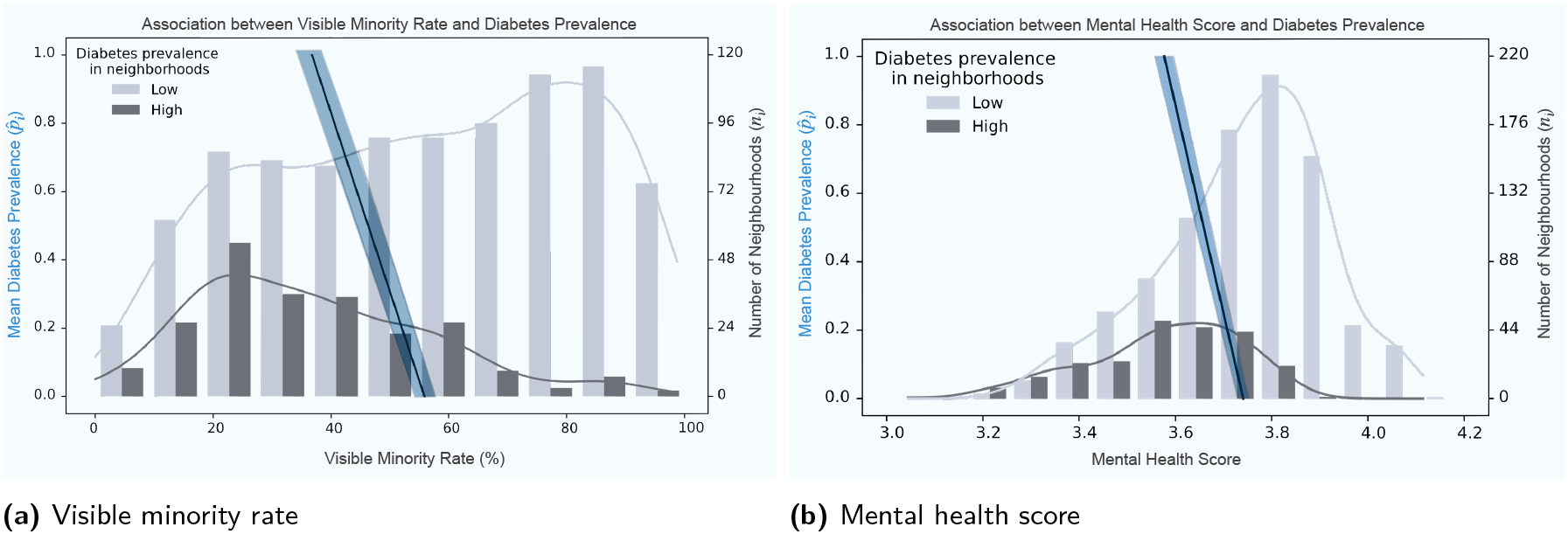
Visible Minority Rates and Mental Health Scores in Relation to Diabetes Prevalence. (a) shows a grouped bar chart with line charts to display binned visible minority rates in neighborhoods classified as having low or high diabetes prevalence. The bar chart uses the right y-axis for the number of neighborhoods *n*_*i*_ in each bin, and the of diabetes presence in neighborhood *k*. (b) applies the same approach to mental health scores. fitted prevalence line uses the left y-axis for the mean diabetes prevalence 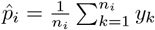, where *y*_*k*_ is a binary indicator of diabetes presence in neighborhood *k*. (b) applies the same approach to mental health scores.

We also found that the `mental health score`, scaled so higher values signify better mental well-being (Table 1), was positively associated with diabetes prevalence. This finding aligns with studies suggesting that compromised mental well-being can raise metabolic risk ^28,29^. As shown in Figure 5b, bins with higher mental health scores show increased mean prevalence, implying that individuals who feel psychologically better may still exhibit biological or lifestyle factors that heighten diabetes risk. The subjective nature of the CCHS measure could produce variability across cultural or socioeconomic contexts ^79^. Non-response bias might inflate this effect if individuals with severe mental distress are less likely to participate, leading to underestimation ^80^. Chronic stress, low mood, and related behaviors can disrupt glycemic control and insulin sensitivity, potentially worsening diabetes risk ^81,82^. Research also points to a feedback loop in which managing diabetes can worsen mental health, creating ongoing adverse metabolic effects ^29^. Supplementary Figures 1 and 2 illustrate how mental health and work stress scores contributed to Toronto CMA models using the extended and full feature sets (Table 4), suggesting that strategies addressing both psychological and metabolic factors could produce enhanced outcomes.

### Causal Forests Insights on Diabetes Risk

Our Causal Forest analysis shows that several variables strongly affect neighborhood-level diabetes prevalence. Work stress has the largest positive effect, with *τ* (*x*) = 0.312. A positive *τ* (*x*) means that rising work stress in neighborhoods with characteristics *x* leads to higher diabetes prevalence, consistent with research linking psychosocial strain to metabolic dysregulation ^83^. Daily smoking rates show a moderate positive influence (*τ* (*x*) = 0.155), aligning with evidence that cigarette use induces inflammation and reduces insulin sensitivity. Mental health scores, however, offer a protective effect: *τ* (*x*) values of *−*1.113 and *−*1.045 in the extended and full feature sets suggest that stronger psychological well-being can lower local diabetes risks.

SHAP analyses support these findings by measuring how each feature influences model predictions. Although active transportation shows minimal causal effect (*τ* (*x*) *≈* 0) in our Causal Forest model, it remains significant in SHAP summaries, possibly reflecting indirect factors such as neighborhood design or added benefits from physical activity. Log median age consistently appears among top predictors, indicating that older populations have higher metabolic risks. Obesity is consistently identified as a key variable with large SHAP values in various analyses, often ranking among the most impactful features. Interestingly, the interplay between work stress and overweight seems stronger than between work stress and obesity, as suggested by nuanced SHAP value patterns. This implies that overweight individuals may have a higher risk of stress-related diabetes, possibly due to different metabolic mechanisms ^84^.

Causal inference combined with ML interpretability clarifies multiple interconnected pathways, emphasizing mental health, stress management, and lifestyle adaptations as important factors in community-level diabetes prevention ^85^. Supplementary Figures 3 and 4 expand on the base, extended, and full feature sets (Table 4), drawing on SHAP-based interpretability and causal inference to explore how psychosocial and demographic attributes shape neighborhood-level diabetes risk.

### Limitations

Although we included a broad range of socioeconomic and behavioral variables, some environmental factors remain unobserved. For example, green space coverage and air pollution, both known to affect health ^86^, are missing from our analysis. We partially addressed this gap using proxies like active transportation and material deprivation indices. However, direct measurements could improve ecological inference, as our Causal Forest model leaves some diabetes risk unexplained. Specifically, while the model estimates *τ* (*x*) for key predictors, an unexplained residual term 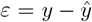 persists, representing unobserved environmental factors or latent confounders. This residual uncertainty suggests that direct measures, such as air pollution levels and green space availability, would enhance our analysis. Moreover, census tracts, despite standardized data collection, may not capture the finer social or mobility patterns that shape neighborhood-level risk ^87^.

Many of our inputs rely on self-reported surveys, raising concerns about recall bias and varying question interpretation ^88^. Although cross-checking with complementary datasets helped reduce these issues, objective clinical assessments could refine model fidelity. The crosssectional design also constrains causal inferences, even when using approaches like Causal Forests ^89–92^. Longitudinal or quasi-experimental studies would offer a clearer view of how social and environmental shifts affect diabetes rates over time.

Our validation strategy, while including external subsets, was limited to the Toronto CMA and City of Toronto, which may narrow generalizability ^93^. Expanding validation to more diverse urban and rural contexts could enhance confidence in model robustness. Due to data constraints, we did not include certain domain-specific variables (e.g., local health services or cultural dietary norms), which may affect broader applicability. While we incorporated several demographic variables, some population segments could remain underrepresented ^94^. Future efforts should include richer data on health, income, and housing in line with recommendations for comprehensive and equitable machine learning in healthcare ^95^.

## CONCLUSION

This study presents a framework merging machine learning classifiers with Causal Forests to identify community factors affecting neighborhood-level diabetes risk in the Toronto CMA. Variables related to physical activity, weight status, and psychosocial stress showed strong ties to local disease patterns, stressing the importance of environmental and behavioral contexts. Combining predictive modeling with causal inference highlighted how certain stress-related and mental health elements might raise or lower diabetes rates at the neighborhood level.

Future research could apply this approach to broader regions and integrate more comprehensive datasets, including longitudinal data and various geographic units. Linking place-based analyses with clinical indicators, healthcare coverage, and dynamic study designs can clarify how neighborhood conditions and social factors change alongside chronic disease outcomes. Enhanced methods may guide public health agencies toward targeted interventions addressing localized needs and promoting long-term well-being among diverse populations.

## ACKNOWLEDGMENTS

This research is supported by the Vector Scholarship in Artificial Intelligence from the Vector Institute, the Artificial Intelligence for Public Health (AI4PH) Trainee Award funded by the Canadian Institutes of Health Research (CIHR).

## AUTHOR CONTRIBUTIONS

Mohammad Noaeen, Amirhossein Rostami, and Ibrahim Ghanem jointly designed and implemented the machine learning pipeline, conducted the data analysis, and drafted the manuscript. Mohammad Noaeen and Amirhossein Rostami led the interpretation and analysis of results by developing the causal ML framework and creating visualizations. Amirhossein Rostami contributed to the web platform development. Zahra Shakeri provided supervision and contributed to data visualization. Olli Saarela, Karim Keshavjee, Jeffrey R. Brook, and Zahra Shakeri provided critical feedback throughout the revision process. All authors reviewed and approved the final manuscript.

## SUPPLEMENTARY MATERIAL

Supplementary material is available at Journal of the American Medical Informatics Association online.

## DATA AVAILABILITY

All databases used in this study are standard, publicly available databases.

## SUPPLEMENTARY INFORMATION

### 1 SUPPLEMENTARY NOTE 1: SENSITIVITY ANALYSIS FOR THE EXTENDED MODELS

**Supplementary Figure 1.**
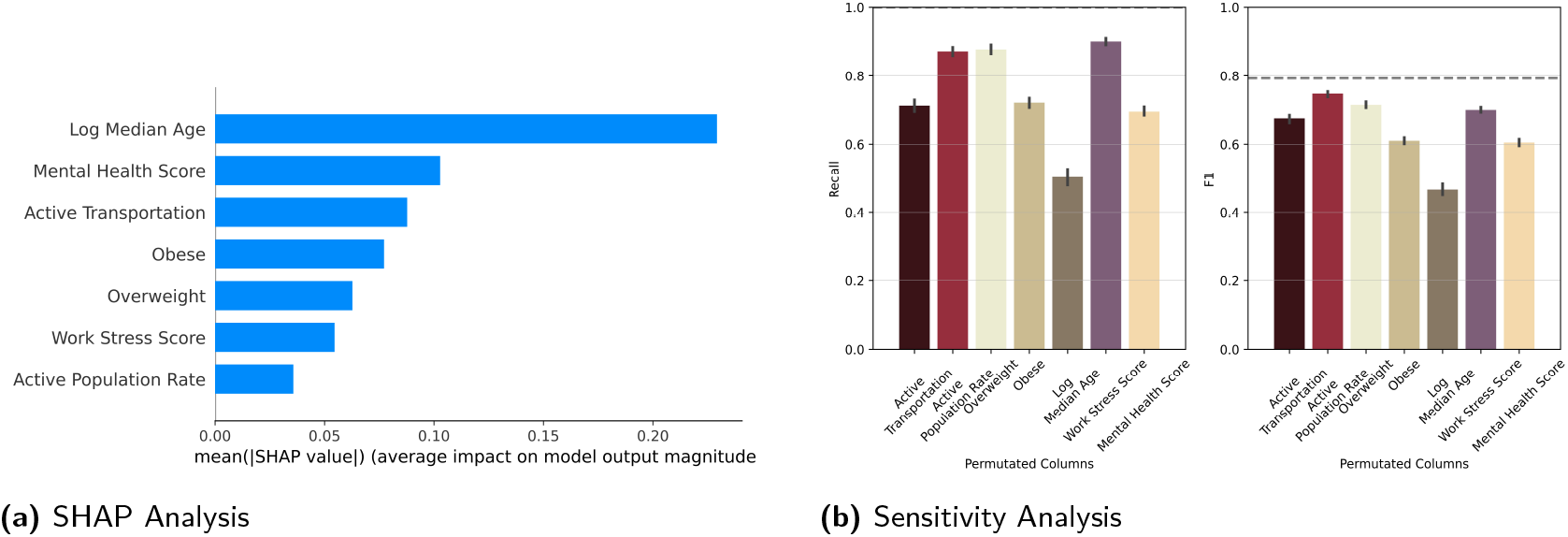
SHAP and Sensitivity Analysis of the extended SVM model on the Toronto CMA dataset with seven variables (Test dataset). (a) shows that Mental Health Score emerges as a highly important feature, with strong overlap in the Sensitivity Analysis. While not perfectly aligned, both analyses rank Mental Health Score as the second most important feature, whereas Work Stress Score appears second least important. (b) indicates that among the two added features, Mental Health Score has the second largest impact, while Work Stress Score shows the lowest effect on the model’s sensitivity. The figure presents the most reliable extracted result from multiple runs.

#### 1.1 Extended SHAP and Sensitivity Analysis for the SVM Model (Test Dataset)

Supplementary Figure 1 presents the outcomes of the extended analysis for the SVM model on the Toronto CMA dataset, using seven variables in the Test set: (a) shows that `Mental Health Score` has high SHAP values, suggesting a strong influence on diabetes risk. This is consistent with the SHAP formula,

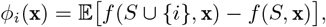

which calculates the local effect of each feature by comparing predictions with and without that feature across all subsets *S*. Mental well-being may contribute to or protect against diabetes risk through stress-related metabolic changes or lifestyle patterns; (b) confirms that permuting Mental Health Score leads to the largest drop in recall and F1, reinforcing its central role in identifying higher-prevalence neighborhoods. Work Stress Score shows a lesser impact, possibly because it captures a narrower range of psychosocial influences compared to overall mental well-being.

These outcomes support the idea that mental health interventions could help reduce diabetes prevalence in at-risk communities, since psychosocial factors appear critical at the neighborhood level. The figure reflects the most stable result among multiple runs, highlighting that both local contributions (SHAP) and global feature sensitivity (permutation test) point to the importance of addressing psychological and behavioral factors in diabetes risk estimation.

#### 1.2 Extended SHAP and Sensitivity Analysis for the SVM Model (External Dataset)

Supplementary Figure 2 presents a parallel analysis for the external dataset: (a) again identifies Mental Health Score as a key predictor, ranking it second only to Log Median Age. This result points to a possible paradox in real-world settings. Certain communities may report strong social ties or cultural practices that elevate self-reported mental well-being, yet those same areas might lack safe venues for physical activity or face limited access to healthy foods, pushing diabetes risk upward; (b) shows that permuting Mental Health Score causes a marked drop in recall and F1, while Work Stress Score has the smallest impact. These observations imply that improving mental health alone may not be enough if local economic or infrastructural barriers remain, suggesting a complex interaction between psychosocial and environmental factors in shaping neighborhood-level diabetes outcomes.

**Supplementary Figure 2.**
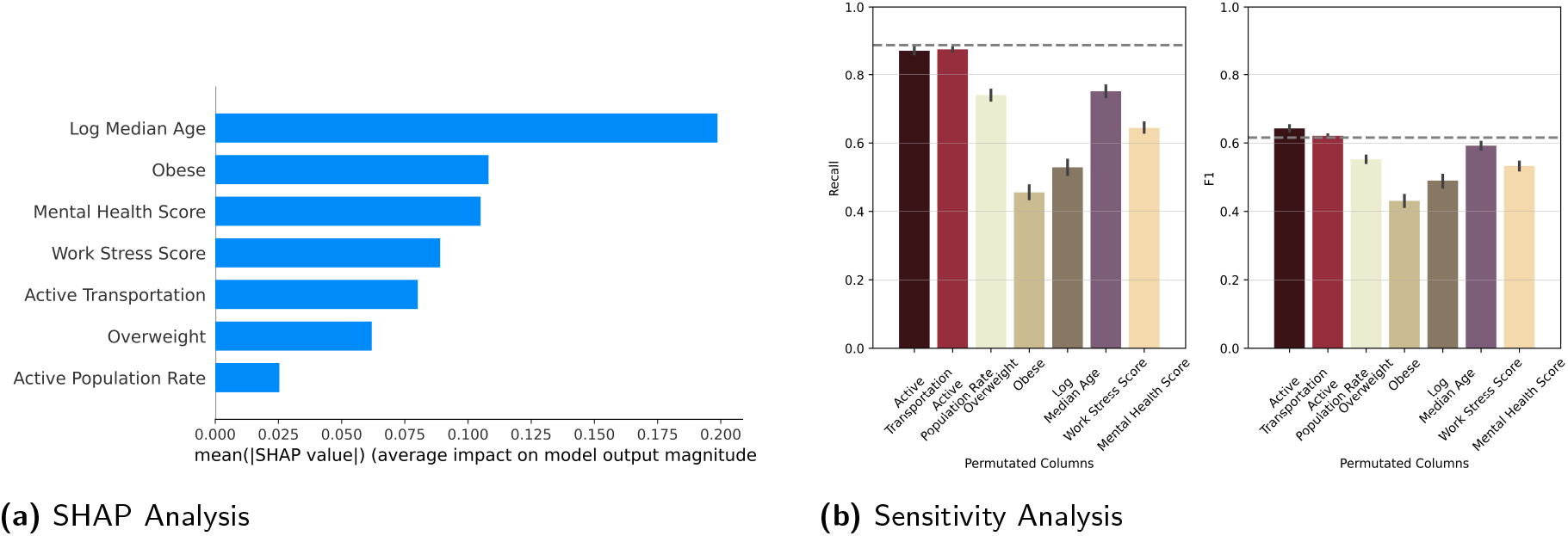
SHAP and Sensitivity Analysis of the extended SVM model on the Toronto CMA dataset with seven variables (External dataset). (a) demonstrates that Mental Health Score emerges as a highly important feature, consistently ranking as the second most influential variable while Work Stress Score ranks as the second least important. (b) shows that among the two added features, Mental Health Score has the second largest impact on model sensitivity, whereas Work Stress Score exhibits the smallest effect. The figure presents the most reliable result obtained from multiple runs.

### 2 SUPPLEMENTARY NOTE 2: ITERATIVE SHAP VISUALIZATION FOR SVM FEATURE SETS

#### Iterative SHAP Visualization for SVM Feature Sets

Supplementary Figure 3 presents the comparison of the SVM model’s base feature set (with five variables) and an extended set (with nine variables): Figure 3a includes Active Transportation and Rate of Active Population as potential treatment variables, whileFigure 3b adds elements such as Mental Health Score, Work Stress Score, and Visible Minority Rate. These additions provide a broader view of the social and behavioral influences that might shape neighborhood-level diabetes risk. Each panel reports SHAP values for selected treatment features, with the remaining ones considered covariates, clarifying how each variable contributes to the model’s predictions under different configurations. Log Median Age, `Obesity`, and `Overweight` are excluded from the treatment sets to focus on psychosocial and mobility-related factors.

This iterative approach clarifies how adding new variables can shift the apparent importance of certain predictors. `Active Transportation` consistently shows strong contributions in both panels, though the introduction of `Mental Health Score` and `Visible Minority Rate` modifies the relative influence of `Rate of Active Population`. These insights go beyond the simple ranking of features, indicating that psychosocial, demographic, and behavioral factors interact in complex ways when shaping diabetes risk. The figure thus provides a richer perspective on how incremental expansions of the feature space can refine understanding of community-level health patterns, suggesting that targeted interventions may need to address multiple domains simultaneously.

**Supplementary Figure 3.**
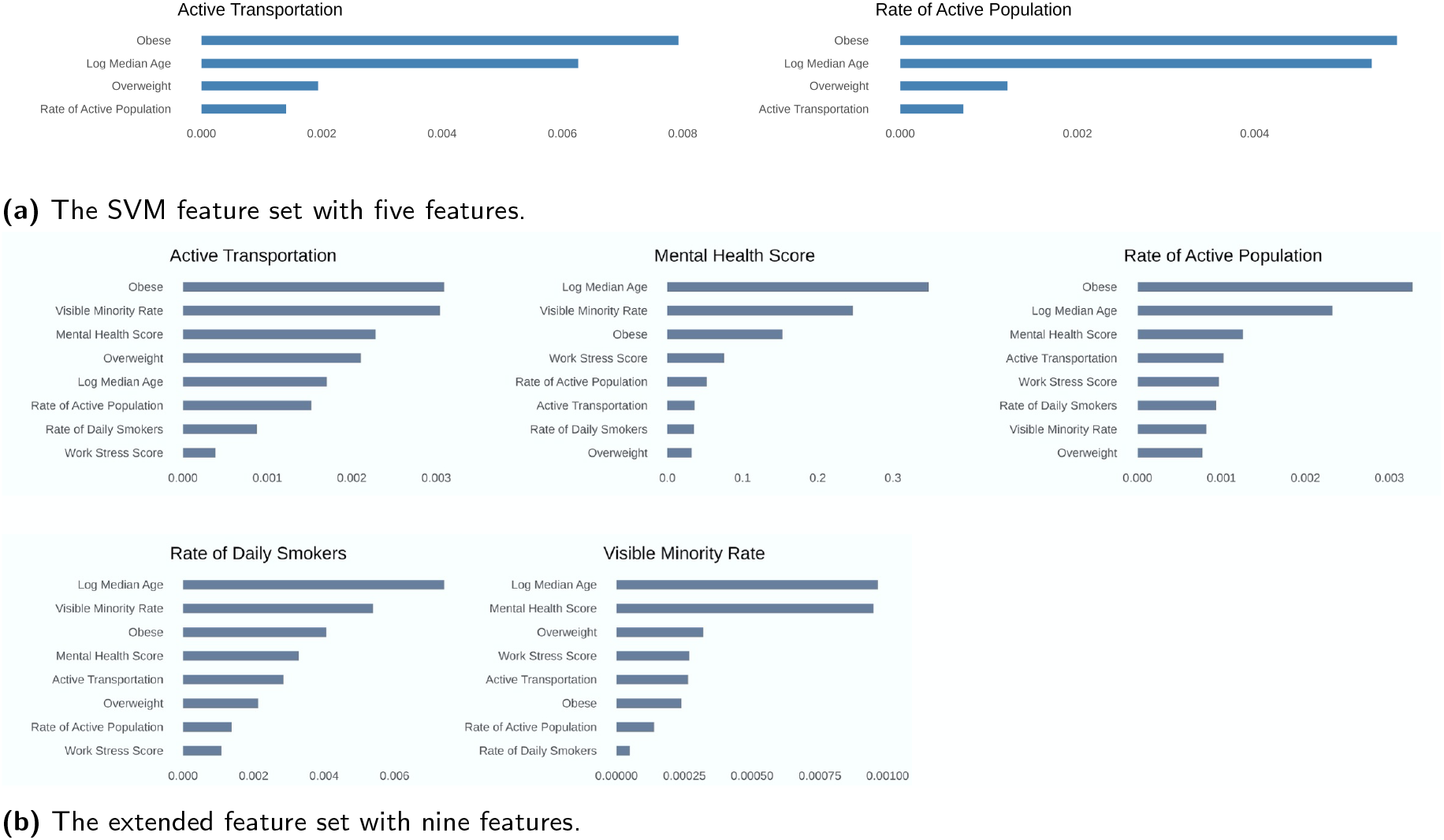
Iterative SHAP visualization: Analyzing feature impact by selecting one as treatment and others as covariates in a) the SVM feature set, and b) the extended feature set. Log median age, Obesity, and Overweight are excluded from the treatment sets.

#### 2.2 Understanding Feature Interactions at Scale

Supplementary Figure 4 provides a detailed view of the full feature set for the SVM model, examining how each variable influences neighborhood-level diabetes risk when designated as the treatment while the rest act as covariates. The sub-figures highlight `Active Transportatio`n, `Average Income`, `High Education Rate`, `Mental Health Score`, `Rate of Daily Smokers`, `Rate of Regular Alcohol`, `Food Insecurity Score`, `Unemployment Rate`, `Rate of Active Population`, and `Work Stress Score`. The expanded selection of variables indicates that psychosocial and socioeconomic factors may overlap in their effects, at times obscuring simpler associations observed in smaller feature sets. For instance, `Work Stress Score` can appear less influential in the presence of `Mental Health Score`, and `Average Income` may combine with `High Education Rate` in ways that affect local prevalence. The figure suggests that diabetes risk arises from a complex mix of personal habits, resource availability, and social conditions.

**Supplementary Figure 4.**
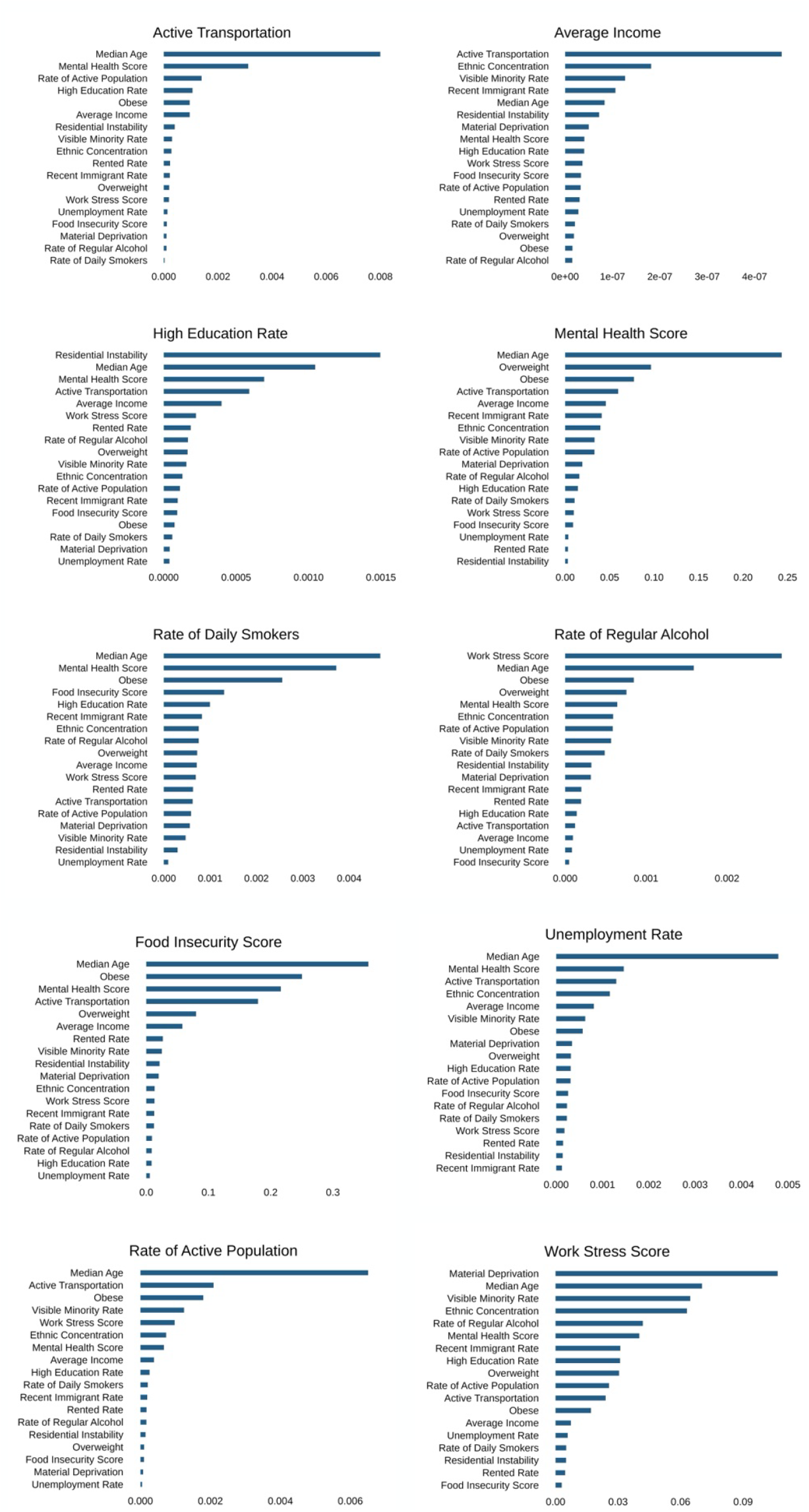
Iterative SHAP visualization: Analyzing feature impact by selecting one as treatment and others as covariates in the full feature set.

#### Footnotes

1 https://github.com/HIVE-UofT/diabetes-analysis

2 https://nediaml.hivelab-uoft.ca/

3 https://github.com/openscilab/pymilo

